# Epidemic Analysis of COVID-19 in Egypt, Qatar and Saudi Arabia using the Generalized SEIR Model

**DOI:** 10.1101/2020.08.19.20178129

**Authors:** Ahmed E. Fahmy, Mohammed M. El-desouky, Ahmed S.A. Mohamed

## Abstract

**Background:** Since its emergence in late December 2019 and its declaration as a global pandemic by World Health Organization (WHO) on March 11, 2020, the novel coronavirus disease known as (COVID-19) has attracted global attention. The process of modeling and predicting the pandemic behavior became crucial as the different states needed accurate predictions to be able to adopt suitable policies to minimize the pressure on their health care systems. Researchers have employed modified variants of classical SIR/SEIR models to describe the dynamics of this pandemic. In this paper, after proven effective in numerous countries, a modified variant of SEIR is implemented to predict the behavior of COVID-19 in Egypt and other countries in the Middle East and North Africa region (MENA).

**Methods:** We built MATLAB simulations to fit the real data of COVID-19 Active, recovered and death Cases in Egypt, Qatar and Saudi Arabia to the modified SEIR model via Nelder-Mead algorithm to be able to estimate the future dynamics of the pandemic.

**Findings:** We estimate several characteristics of COVID-19 future dynamics in Egypt, Qatar and Saudi Arabia. We also estimate that the pandemic will resolve in the countries under investigation in February 2021, January 2021 and 28th August 2020 With total death cases of 9,742, 5,600, 185 and total cases of 187,600, 490,000, 120,000, respectively.

## 1 Introduction

The coronavirus disease 2019 (COVID-19) is a novel viral infection that was first detected in December 2019 in Wuhan, China. It quickly prevailed in different countries all around the world, and it was in March 11 that the WHO declared it a pandemic [1]. Although the disease is one of the family of corona viruses, it has some characteristics that differentiate it from other viruses (like SARS in 2003) such as high infection rates during the incubation period which is reported by [2], as a median, to be 5.1 days with 14 days at maximum, and also as the time between the real dynamics and the confirmation of the infected cases due to the high speed of transmission of this virus. The fact that no symptoms of the disease show at the incubation period contributes to its ability to spread to a large number of people. Known modes of the transmission of COVID-19 are through respiratory outputs (like the droplets) followed by physical contact between individuals or surfaces. As a result, new public safety measures have been implemented to try and mitigate the impact of the pandemic such as teleworking, cancellation of public events, suspending offline education, and closure of clubs and other venues open for social gatherings. Since 30 July 2020, the number of confirmed cases reached 16.81 million with 662,000 deaths around the world [3].

One of the main challenges in studying epidemics is trend prediction of the disease behavior; that is to predict how many individuals will be infected in the future, determining the peak of the pandemic, the possibility of a second wave of the disease and the final number of deaths at the pandemic‘s end. Based on analysis of these trends, the governments take actions to try to limit both human and economic losses as much as possible. Currently, the COVID-19 outbreak is the dominating issue threatening humanity; researchers from different backgrounds - such as applied mathematics, epidemiology, and data science - have been working on studying the outbreak.

The start of epidemic modeling was in 1927 by Kermack and McKendric when they developed models to study the plague and cholera diseases [4, 5]. After that, several models have been developed in order to study the infection of viruses among individuals such as the SIR model [6,7,8,9] which contains 3 categories: Susceptible, Infectious, and Recovered. One of the most interesting models is the SEIR model [10,11,12,13] which contains 4 categories: Susceptible, Infectious, Exposed, and Recovered. The SEIR model can be derived from the SIR model and it requires a large amount of data to give valid results. The SEIR is the most widely used model in investigating and studying COVID-19 in different countries around the world [14]. Since the outbreak started, the SEIR model has been used successfully to assess the measures effectiveness, a task that is not easy via the statistics models. In [15], the SEIR model was used to study the effect of the lock-down in Hubei, China on the transmission rate of COVID-19 in Beijing and Wuhan. Because of the distinction that COVID-19 is transmitted during the latent period, the classical models such as the SIR, SEIR, and SEIJR [16] (which contains 5 categories: susceptible, exposed, infectious, diagnosed, and recovered) models cannot be used effectively to study the outbreak of COVID-19. Nevertheless, taking that point into account, Peng et al. [17] introduced a generalized version of the classical SEIR model, the (SEIQRDP) model, to study the dynamics of the COVID-19 outbreak in China. The generalization introduces new states:quarantined state(Q), death state(D) and protected state (P). It also considers the effects of the preventive decisions on the main parameters such as quarantine time and the latent period. Based on this generalized model, Peng et al. [17] had very successful results on the pandemic dynamics in China. The generalized model was widely adopted in most literature. For example, Al-Hussein et al. [18] used it to study the outbreak in Iraq, Mangoni and Pistilli [19] could analyze the outbreak‘s dynamics through it in Italy. In addition, Bahloul et al. [20] applied the model in China, France, and Italy.

In this paper, we collect the data of COVID-19 in Egypt, Saudi Arabia and Qatar in the period from 5 April to 26 July, and apply the generalized SEIR model to study the epidemiological characteristics of the disease in each of these countries and to understand how the COVID-19 would evolve.

The structure of the paper is as follows. Section 2 describes the employed model and its details. Section 3 discusses the utilized data sets and parameter estimation algorithm. In section 4, the results of the proposed model are illustrated and discussed for each country. Finally, in section 5, we summarize the study and draw concluding remarks.

## 2 Model

As introduced by Peng in [17], the SEIQRDP model consists of seven compartments defined as follows:

*S*(*t*): Number of susceptible cases.
*E*(*t*): Number of exposed cases (infected, but not infectious).
*I*(*t*): Number of Infectious cases, but not quarantined.
*Q*(*t*): Number of quarantined cases.
*R*(*t*): Number of recovered cases.
*D*(*t*): Number of dead cases.
*P* (*t*): Number of insusceptible or protected cases.

There are also six parameters that characterize the dynamics of the disease in each country, they are *{α, β, γ, δ, λ(*t*), κ(*t*)}* and they represent the protection rate, the infection rate, the inverse of the average latent time, the rate of quarantining, the cure rate, and the mortality rate, respectively.

The interaction between the 7 compartments is depicted in figure 1.

**Figure 1:**
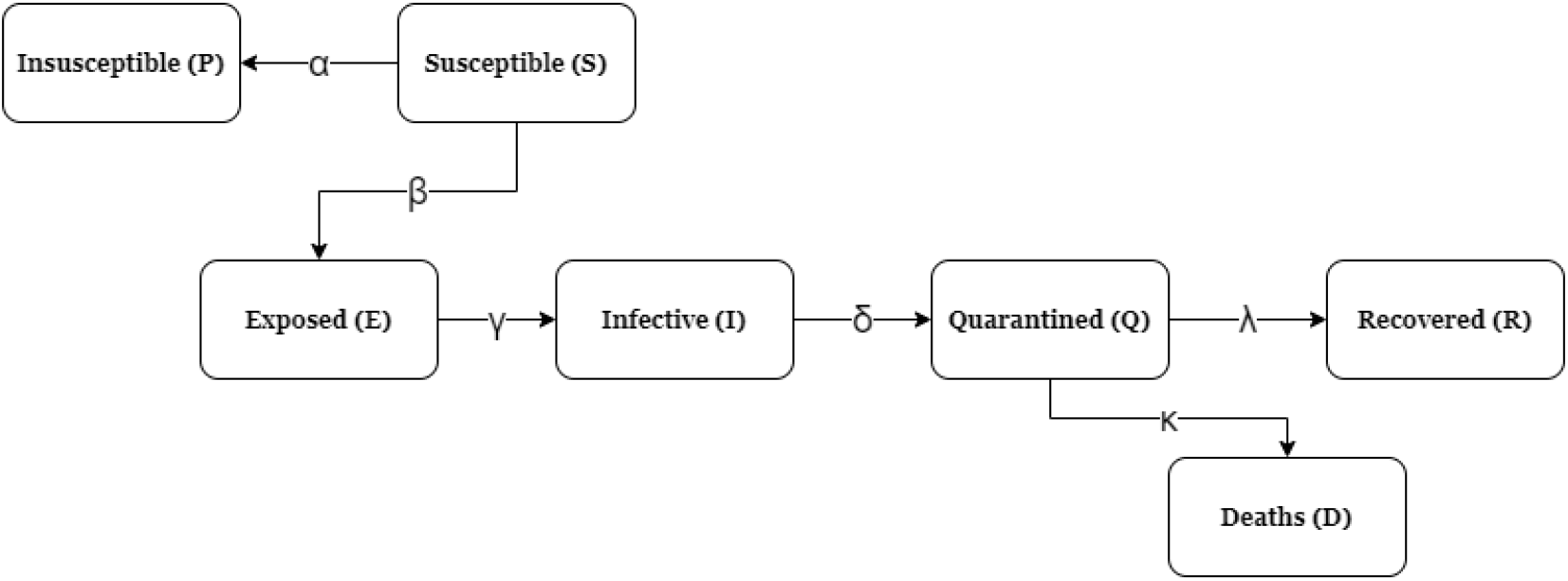
The interactions between the 7 compartments of the generalized SEIR model

The above compartments are related via the following system of nonlinear coupled ODEs:

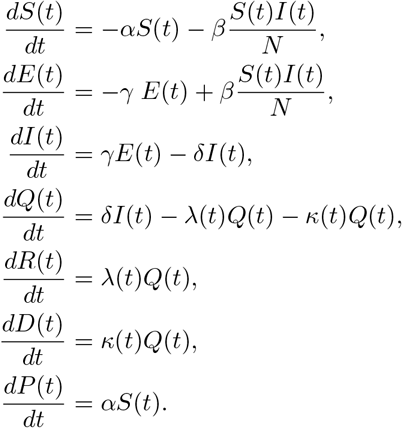

Here, *N* represents the population of the country under study where it satisfies:

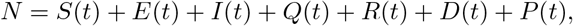

and by direct differentiation with respect to time, we get:

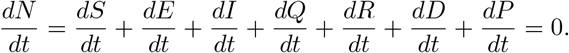

The last two equations are valid under the assumption that the birth rate and the death rate (due to no COVID-19 infection) are very small compared to the changes due to the outbreak.

Both the cure rate *λ* and the mortality rate *κ* are assumed to be time dependent based on the analyzed collected data in some provinces in China [21]:

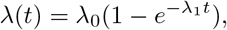

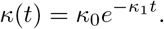

This analysis [21] has shown that the cure rate increases rapidly with time, and the mortality rate decreases rapidly with time, which agrees with common sense. That is, over time people understand the importance of the precautions procedures due to awareness campaigns provided with governmental agencies along with the trials being tested while searching for a drug. It is natural that the cure rate starts at a very small value and then increases till reaching some specific level *λ*_0_ with *λ*_1_ specifies how fast the rate reaches the max limit defined by *λ*_0_, and the opposite occurs for the mortality rate: it starts at some high value *κ*_0_ and decreases with time till reaching zero after a relatively large time with *κ*_1_ characterizing how quick the rate drops to zero. Also, the mortality rate must go to zero as the cure rate reaches its maximum, and that is consistent with any epidemic. Additionally, Peng et al. [17] have confirmed such a behavior when studying some provinces in China.

## 3 Parameter Estimation

The 6 parameters used in the model are key characteristics of the pandemic dynamics that differ drastically between countries [17,22,23]. COVID-19 statistics of countries under investigation were obtained from Johns Hopkins University Center for Systems Science and Engineering [24]. Here, we utilized 8 fitting parameters which are *{α, β, γ, δ, λ*_0_*, λ*_1_*, κ*_0_*, κ*_1_*}*. The fitted parameters are obtained by minimizing the error norm using MATLAB built in nonlinear optimization solver “fminsearch” that employs Nelder-Mead algorithm. The optimized parameters generated by the solver are illustrated in Table 1. To prevent over fitting we used Peng’s approach [17] with initial estimations of parameters consistent with other attempts to model COVID-19 in (MENA) [25,26,27]. A related algorithm is given below.

**Table 1:** The optimized parameters

| Parameter | Optimized parameters for Egypt | Optimized parameters for Qatar | Optimized parameters for Saudi Arabia |
| --- | --- | --- | --- |
| $\alpha$ | 0.0351 | 0.0530 | 0.0025 |
| $\beta$ | 0.3059 | 0.8404 | 0.23767 |
| $\gamma$ | 0.1198 | 0.0716 | 1.7163 |
| $\delta$ | 0.0187 | 0.0862 | 0.1600 |
| $\lambda_0$ | 0.6767 | 4.7972 | 0.0623 |
| $\lambda_1$ | 0.0003 | 0.0002 | 0.0789 |
| $\kappa_0$ | 0.0018 | 0.00008 | 0.0007 |
| $\kappa_1$ | 0.0021 | -0.001 | -0.0001 |

#### Algorithm 1: Estimation of the fitted parameters

Input: *R*:(recovered cases data). *D*: (death cases data). *I*: (confirmed cases data). *t*: (time in units of days). *param*: initial guess of the parameters (based on their physical meaning and being consistent with the literature). *y*_0_: initial values of the 7 compartments. *fit*: the model to be fitted with the real data. Output: *OptPar*: the fitted parameters to the real data. The optimization function: [OptPar] = fminsearch(@fit,param, options, inputs)

The optimized parameters show typical ranges as per comparison with optimized values present in [23]. The behavior of cure rate *λ*_0_ of Qatar is untypical due to the late start date of simulation (April 10th) along with the strong health care system there along with the small number of population there and the massive testing that takes place. The parameter *κ*_1_ best-fitting values in Qatar and Saudi Arabia were negative values of low orders of magnitude (–0.001, –0.0001), respectively, which implies that the death rates in these countries are approximately constant with a slight increase throughout the simulation. These parameter values are consistent with the slight rise of death cases observed in late May (21st until 29th) in Qatar and Saudi Arabia [24]. This observed behavior of death cases may have caused the death rate to start at a slightly lower value at the simulation starting date (April 10th) and increase very slightly to account for the slight rise of death cases in late May. The documented code is available in the Git repository ^2^.

## 4. Results and Discussion

### 4.1 Prediction of time evolution of COVID-19 for Egypt

Results of simulations of COVID-19 in Egypt are illustrated in figures 2, 3, 4, 5.

**Figure 2:**
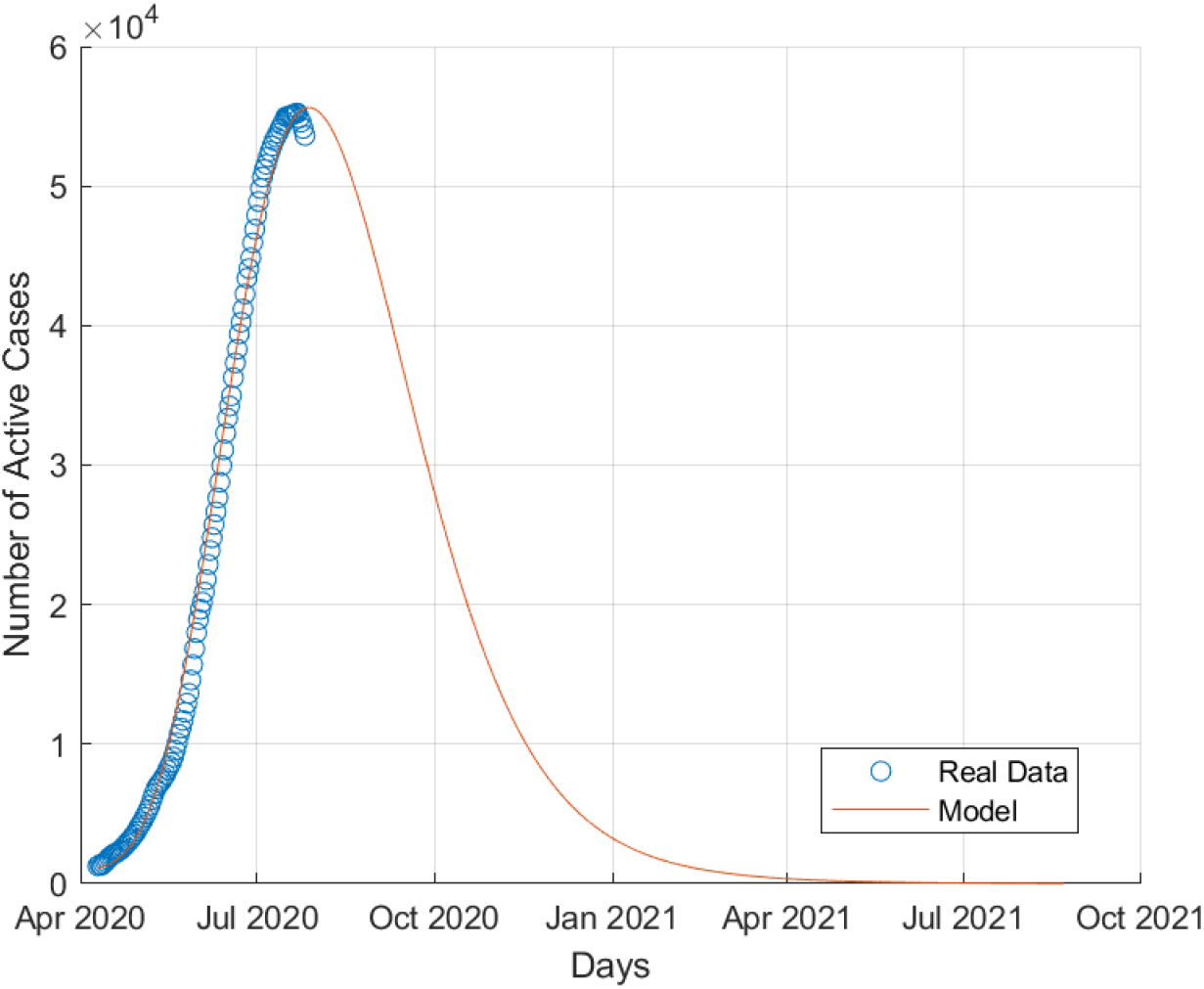
Active COVID-19 cases in Egypt.

**Figure 3:**
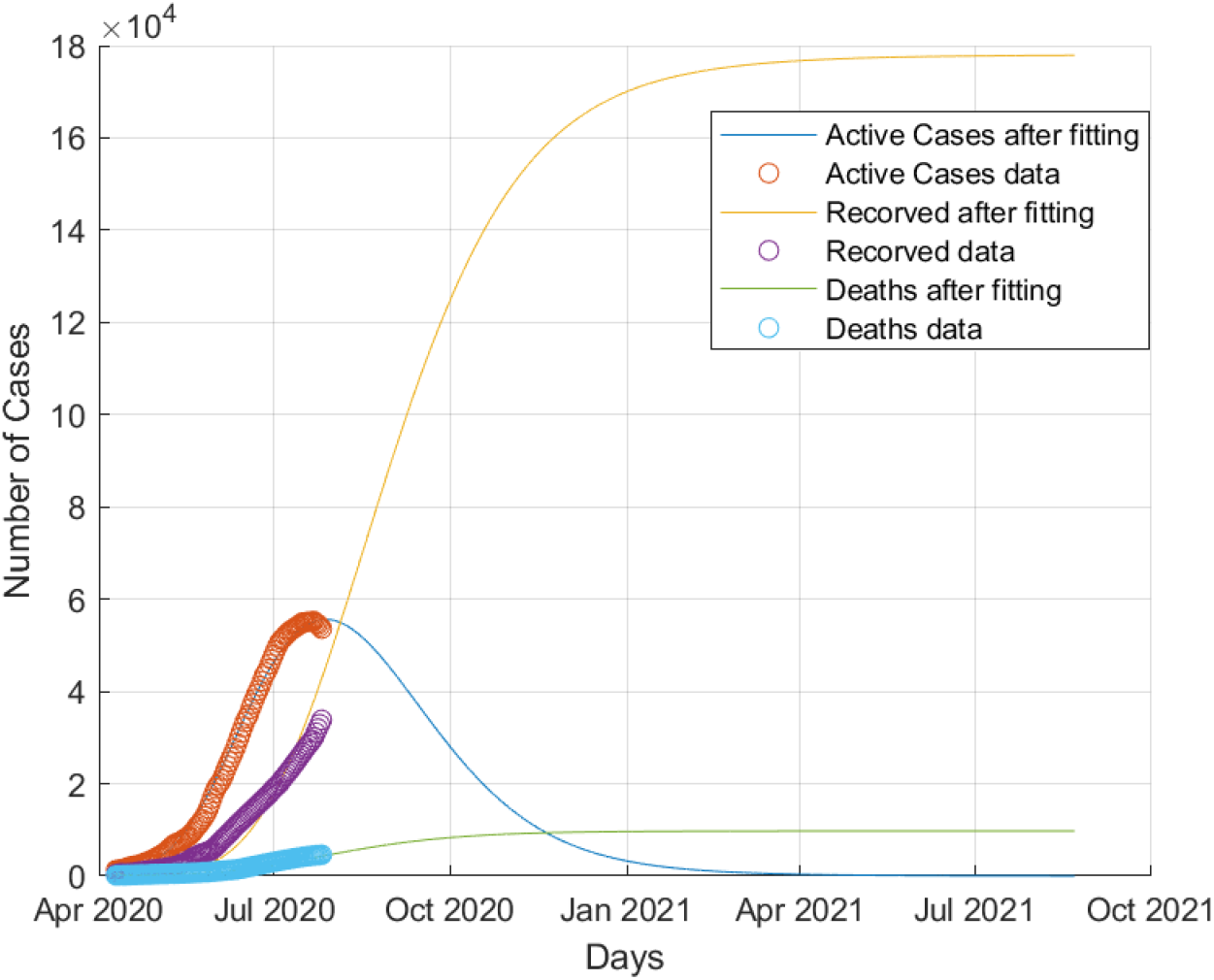
Recovered COVID-19 cases in Egypt.

**Figure 4:**
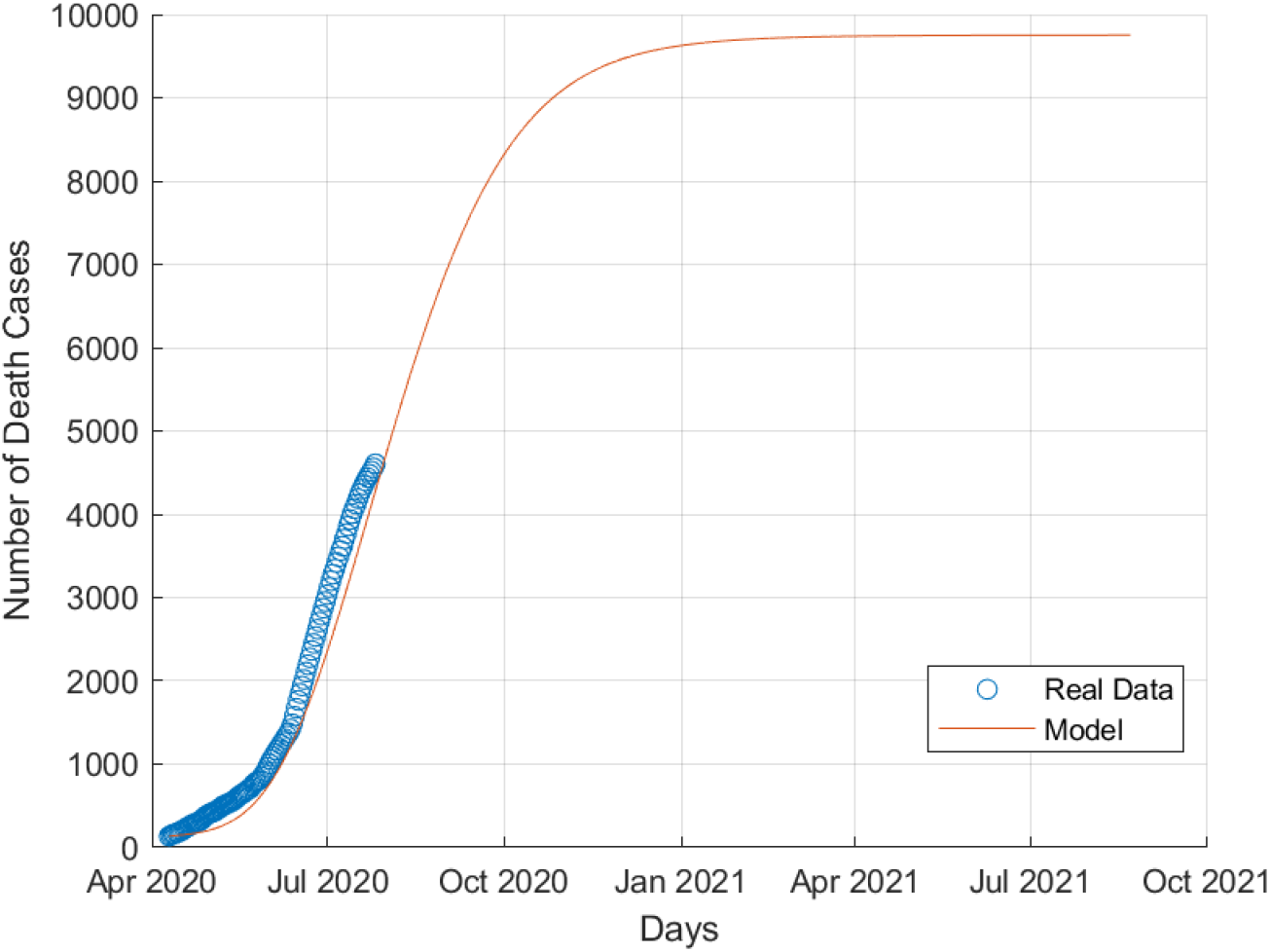
Death COVID-19 cases in Egypt.

**Figure 5:**
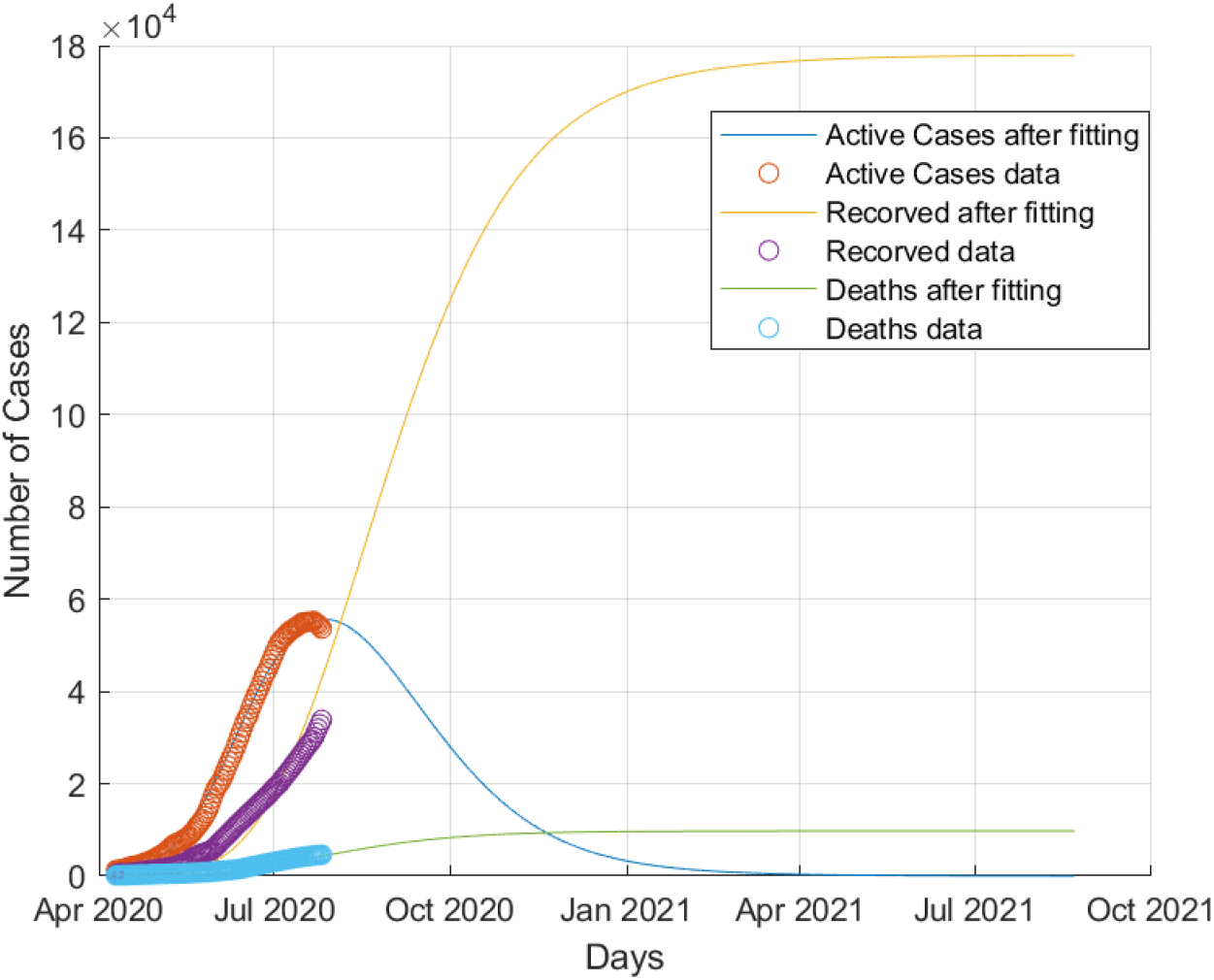
Collective behaviour of COVID-19 dynamics in Egypt.

### 4.2 Prediction of time evolution of COVID-19 for Qatar

Results of simulations of COVID-19 in Qatar are illustrated in figures 6,7,8,9.

**Figure 6:**
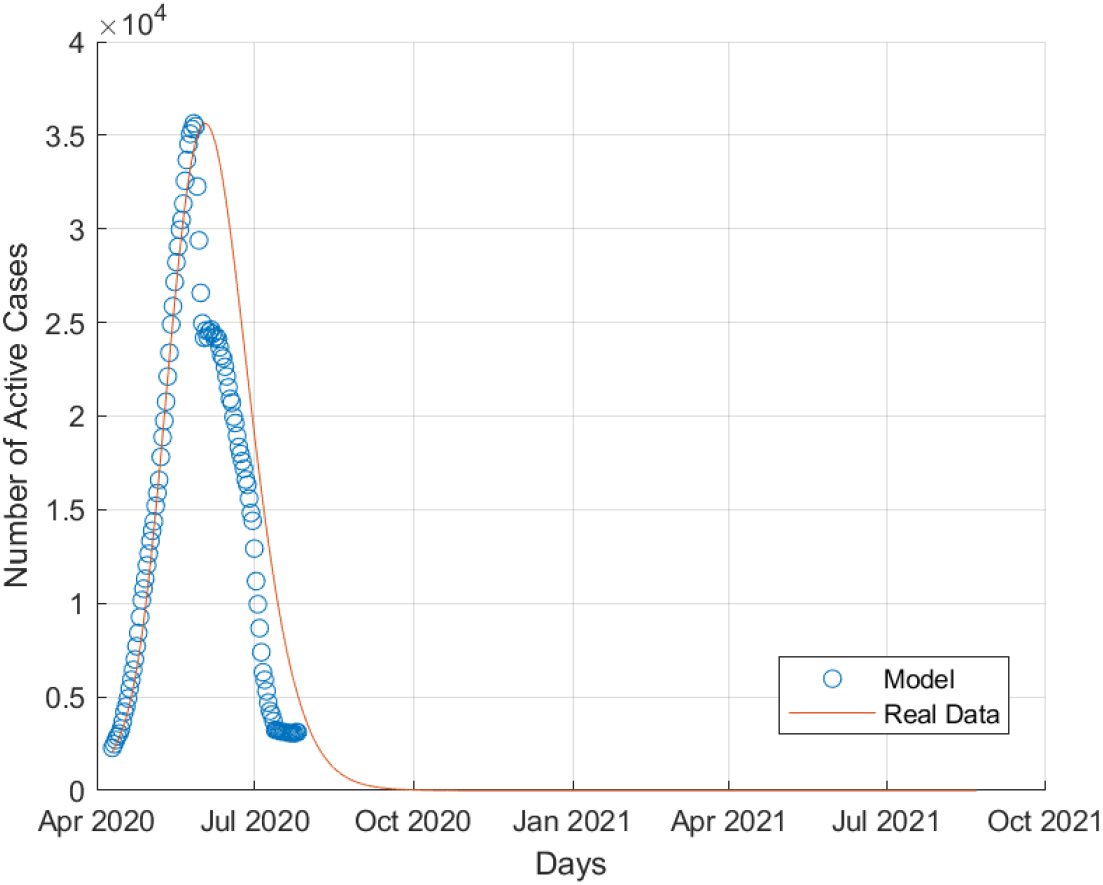
Active COVID-19 cases in Qatar.

**Figure 7:**
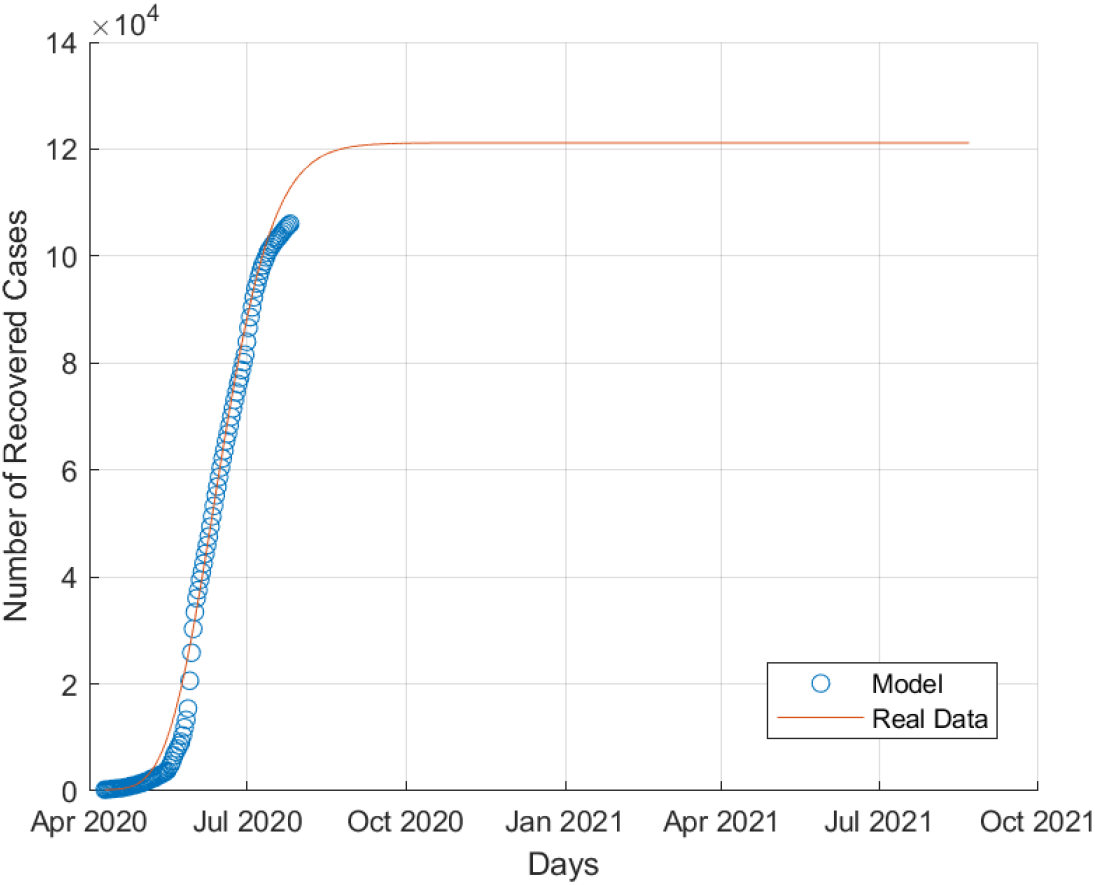
Recovered COVID-19 cases in Qatar.

**Figure 8:**
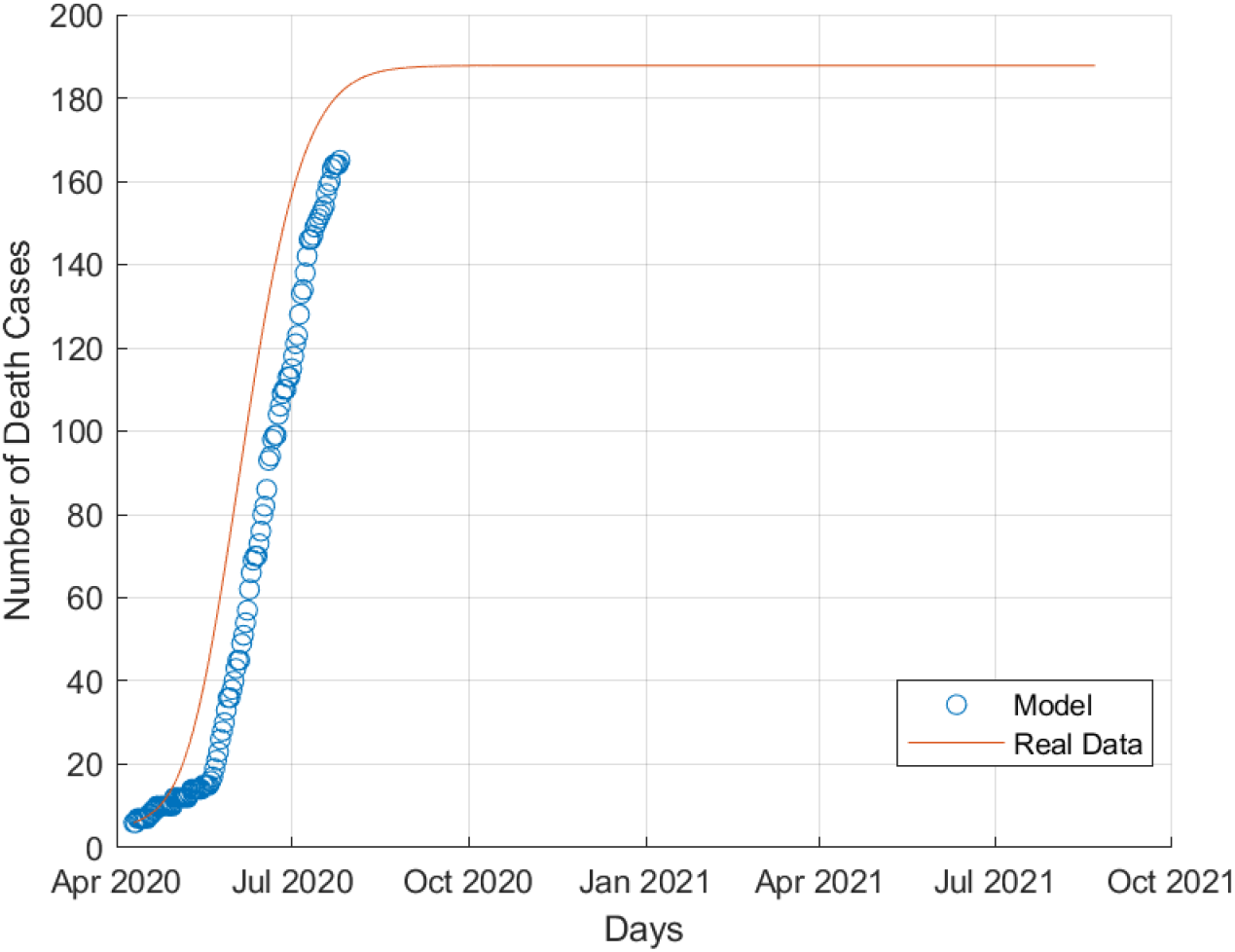
Death COVID-19 cases in Qatar.

**Figure 9:**
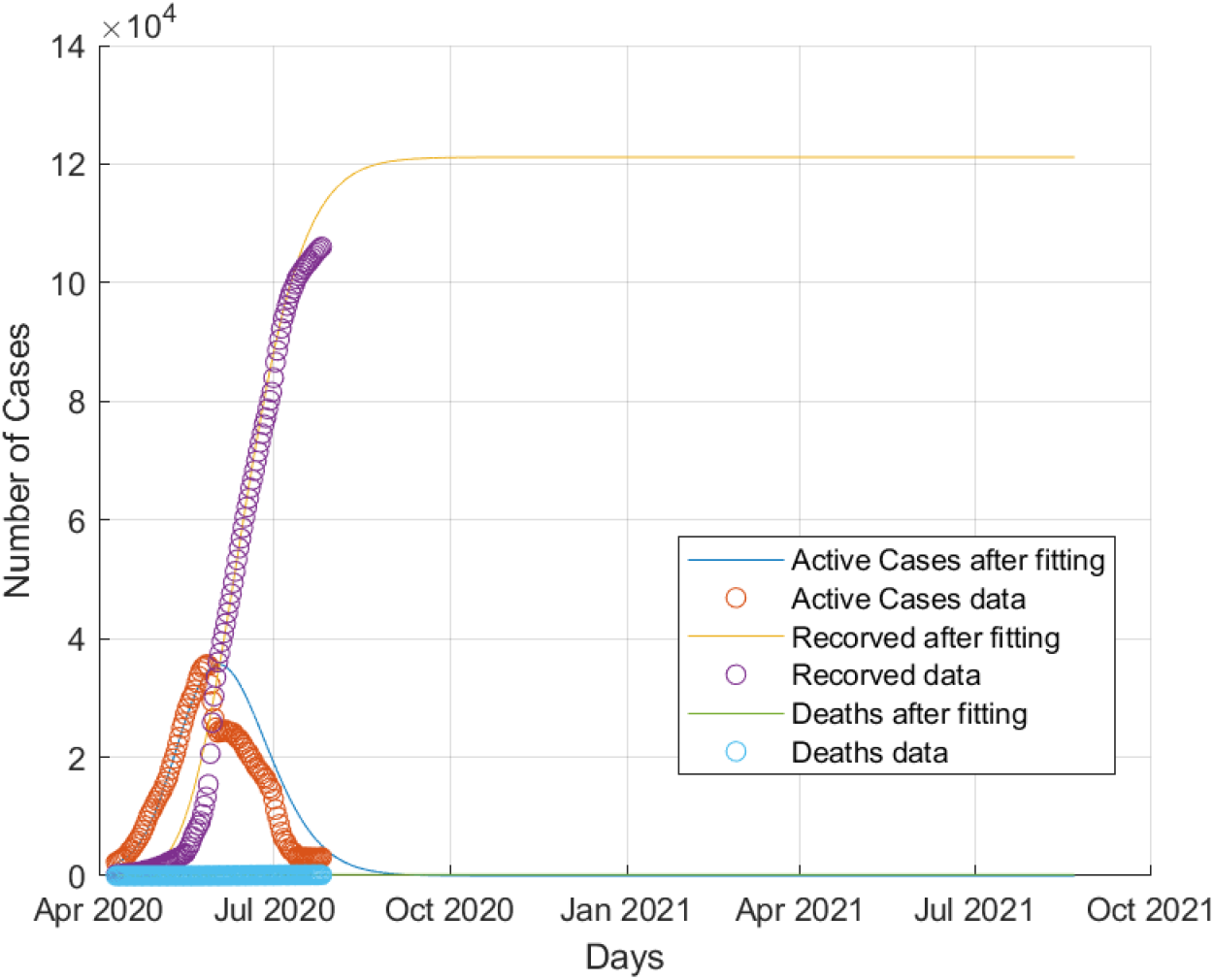
Collective behaviour of COVID-19 dynamics in Qatar.

### 4.3 Prediction of time evolution of COVID-19 for Saudi Arabia

Results of simulations of COVID-19 in Saudi Arabia are illustrated in figures 10,11,12,13.

**Figure 10:**
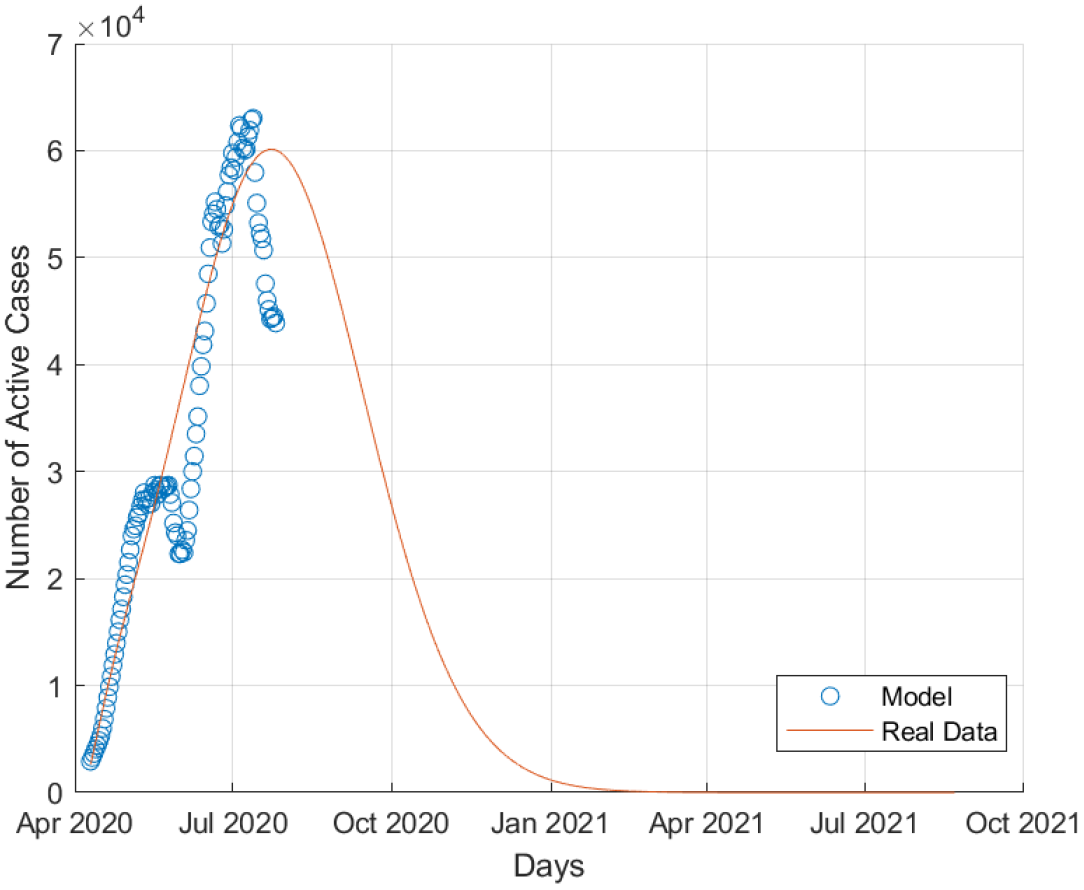
Active COVID-19 cases in Saudi Arabia.

**Figure 11:**
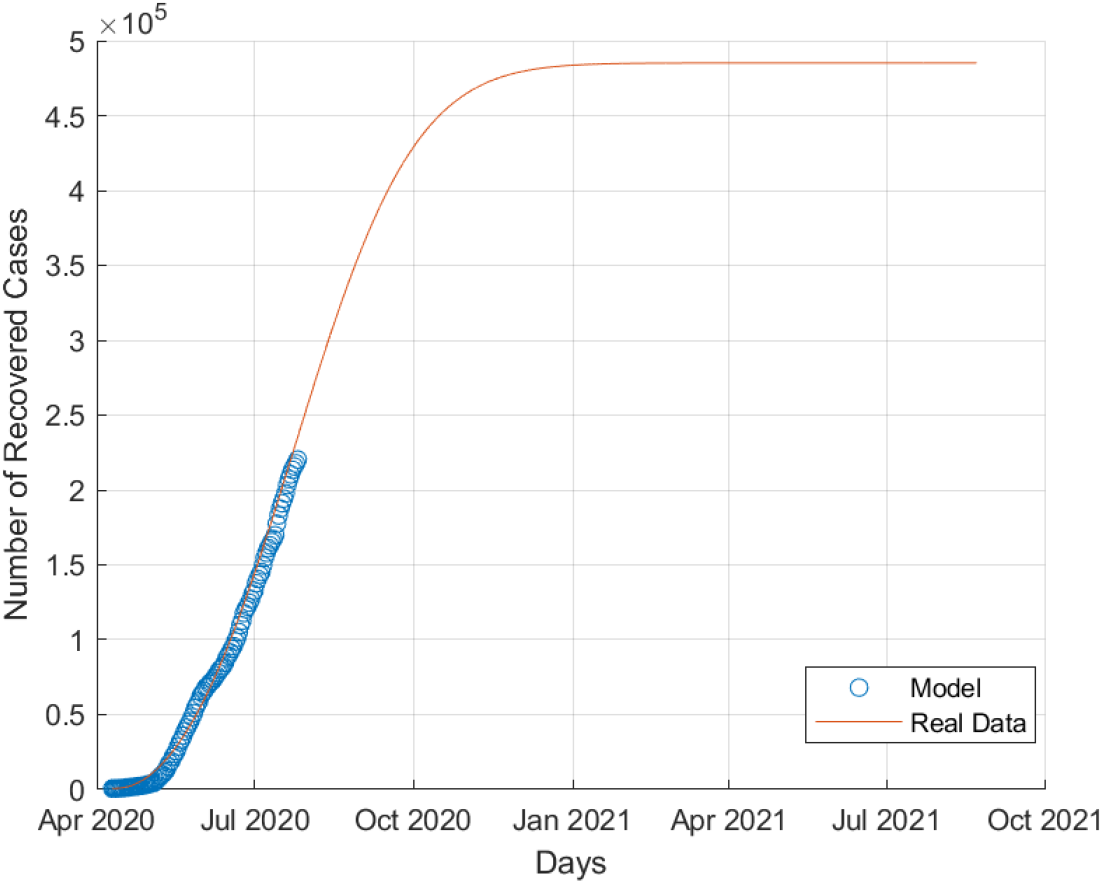
Recovered COVID-19 cases in Saudi Arabia.

**Figure 12:**
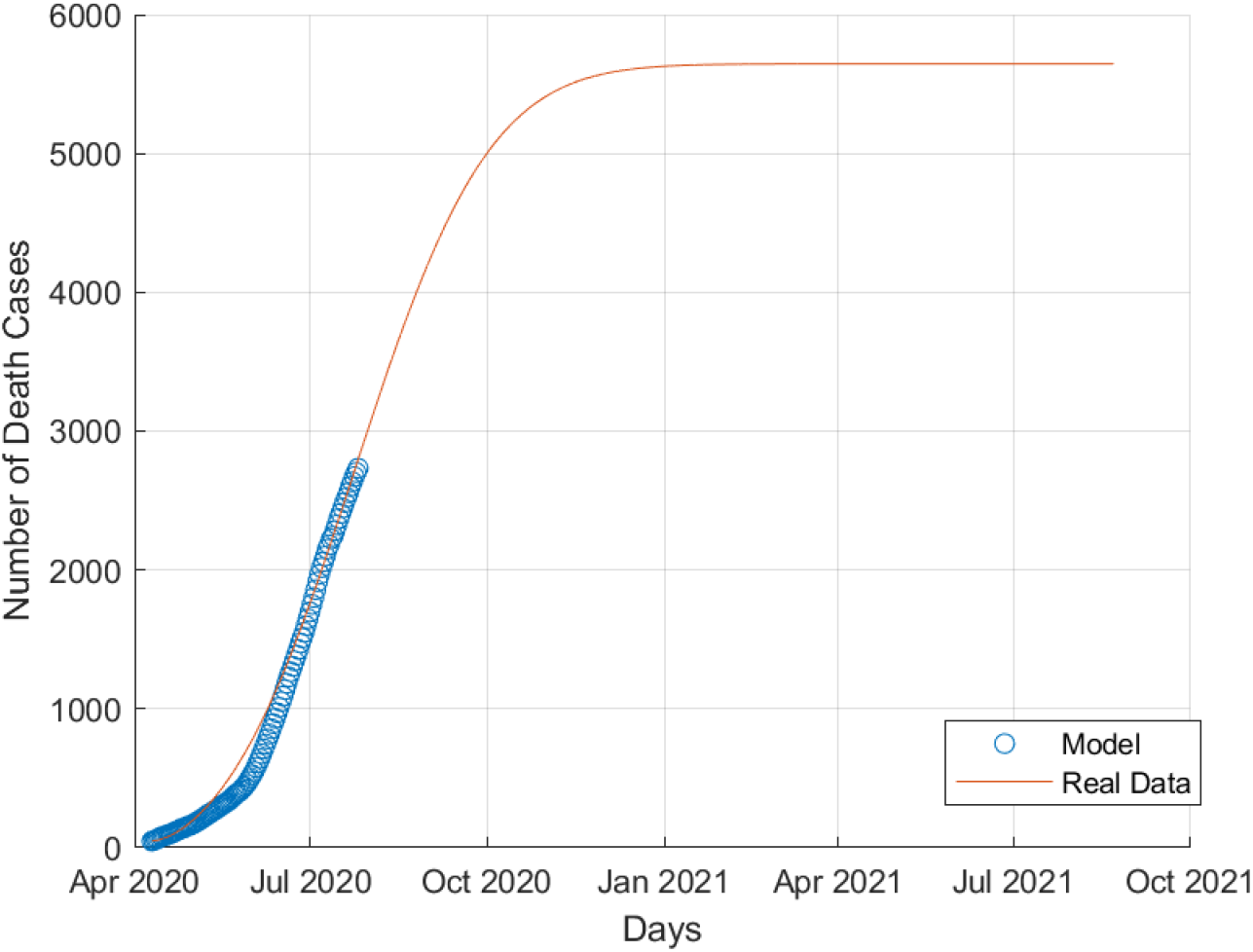
Death COVID-19 cases in Saudi Arabia.

**Figure 13:**
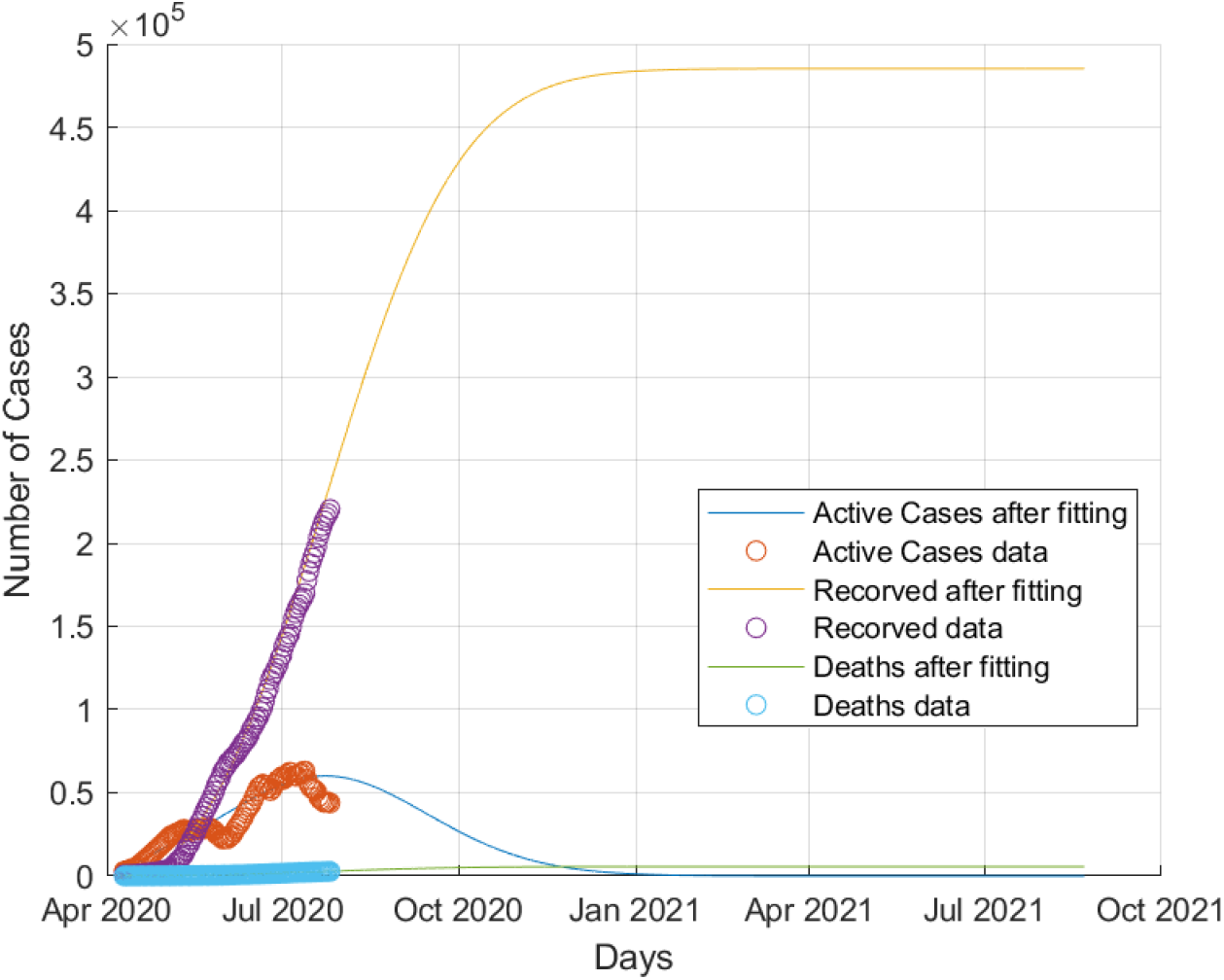
Collective behaviour of COVID-19 dynamics in Saudi Arabia.

### 4.4 Estimation of COVID-19 dynamics in Egypt, Qatar and Saudi Arabia

Assuming no major medical breakthroughs during simulation time or other Non-Pharmaceutical Interventions (NPIs) introduced, Table 2 summarizes resulted simulations based estimations for the time evolution of COVID-19 dynamics in Egypt, Qatar and Saudi Arabia. Here, we employed halving date, as a relative measure of the decay of the number of active cases. Halving date measure represents the date at which the active cases in the country under investigation have declined into half of its value recorded at the last data point of the COVID-19 data sets used in running simulations (26th of July 2020). These recorded values of active cases on the 26th of July 2020 are 53,630, 3,116, 43,390 for Egypt, Qatar, Saudi Arabia, respectively.

**Table 2:** Estimations for the time evolution of COVID-19 dynamics in Egypt, Qatar and Saudi Arabia

| **Country** | **Total Deaths Estimation** | **Total Cases Estimation** | **Active Cases Halving Date Estimation** | **Peak Active Cases Date Estimation** | **Peak Active Cases Estimation** | **Pandemic Ending Date Estimation** |
| --- | --- | --- | --- | --- | --- | --- |
| **Egypt** | 9742 Cases | 187600 Cases | first week in October 2020 | 22<sup>nd</sup> of July 2020* | 55230 Cases* | February 2021 |
| **Saudi Arabia** | 5600 Cases | 490000 Cases | 8th of September 2020 | 13<sup>th</sup> of July 2020* | 63030 Cases* | January 2021 |
| **Qatar** | 185 Cases | 120000 Cases | 12<sup>th</sup> of August 2020 | 30<sup>th</sup> of May 2020* | 35480 Cases* | 28<sup>th</sup> August 2020 |

The countries denoted by (*) in their “peak active cases date estimation” and “peak number of active cases estimation” cells have already passed the active cases peak phase. Therefore, instead of using estimated values, real values are used.

As illustrated in Table 2, the estimated halving dates are close.

The highest estimated final burden of COVID-19 cases in the countries under investigation is 490,000 cases estimated for Saudi Arabia. The relatively high estimated number of total cases in Saudi Arabia is consistent with their policy of massive testing [28].

When the simulations start date is set earlier to include the period before the application of NPIs and lockdowns, the infectivity rate *β* becomes drastically higher which implies these measures’ importance. Consider the following case study, Egypt started partial lockdown since 15th of March 2020 [29] after adding the incubation period of 5 days as reported in [30], the pandemic dynamics is expected to have changed after 20th of March approximately. When the simulation starting date is set to the 29th of February, the simulation includes 21 daily data points of the non-mitigated pandemic dynamics before lockdown (representing 15% of the length of the data set), the infectivity rate *β* optimized value becomes 0.66 instead of the simulated above the value of 0.3059. Figure 14 shows the behavior of the active cases when the data points before the lockdown are incorporated into the dataset. This behavior is consistent with our current understanding of how NPIs affect COVID-19 dynamics [31]. The same response of COVID-19 dynamics to lockdowns is observed in Saudi Arabia active cases shown in figure 10. Saudi Arabia exhibits double peak behavior in the active cases due to the removal and reapplication of lockdown separated by short period [32].

**Figure 14:**
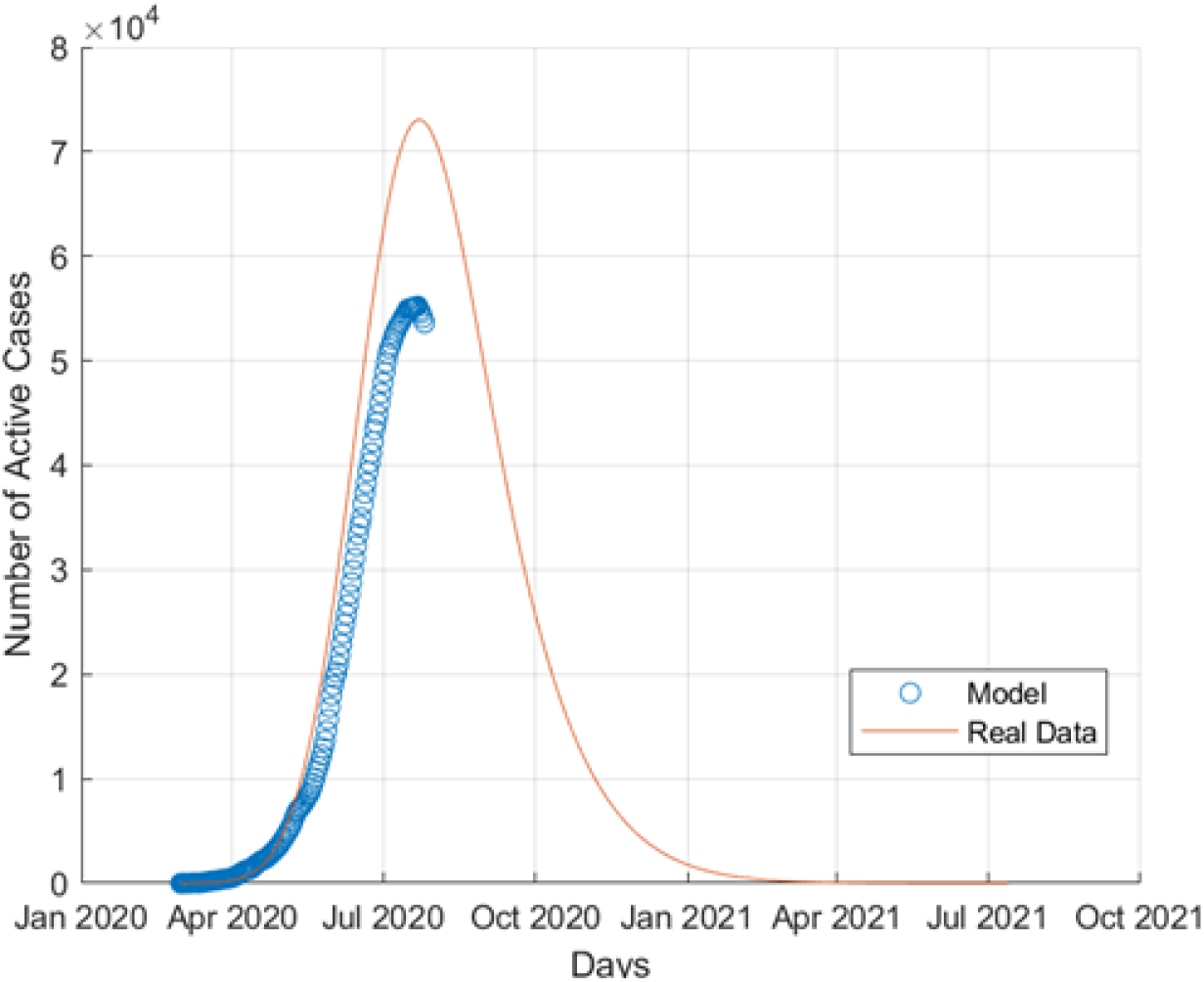
Egypt’s active COVID-19 cases with pre-lockdown data incorporated

## 5. Conclusion

In this paper, we employed a generalized and modified version of classical SEIR model to analyze the dynamics of COVID-19 in Egypt, Qatar and Saudi Arabia. We utilized the MATLAB built-in nonlinear optimization solver “fminsearch” that employs Nelder-Mead algorithm to find the optimized parameters for each country. The system of differential equations are solved numerically to obtain the time evolution of the different mutually exclusive categories of populations suggested by the model: *S, E, I, Q, R, D, P*. We then used these time series to estimate the total number of infected cases and deaths when the active cases become zero in countries under this study. As seen in literature and as provided in the presented results, the generalized SEIR model works effectively in modeling the outbreak and analyzing its dynamics. However, it does not predict a second wave of the current pandemic; a point that will be more clear in the coming few months. If there exists a second wave due to return of the social activities with precautions procedures being neglected, this will need further investigation and will be considered in our future work.

## Data Availability

All the data used in this study are reported by World Health Organization (WHO) and it is publicly available.

## 6. Declaration of Interests

Authors declare no competing interests.

## 7. Funding

This study was NOT funded by any institution.

## 8. Acknowledgments

Authors would like to thank Abdelrahman Abbas and Mohamed Mousa, Zewail City of Science and Technology, Egypt, for their useful discussions.

## 9. Contributions

Both Ahmed Fahmy, and Mohammed El-Desouky contributed equally to this work under the supervision of Ahmed S.A. Mohamed.

## Footnotes

2 https://github.com/AhmedFahmy177/Generalized-SEIR-model-for-COVID-19-Modeling.git

## Notes

### Competing Interest Statement

The authors have declared no competing interest.

### Author Declarations

NO IRB/oversight body is needed in this study.

